# Association between TikTok use and the development of eating disorders in young Colombian adults between 18 and 25 years of age

**DOI:** 10.1101/2024.10.22.24315901

**Authors:** Laura Restrepo-Escudero, Silvia McCormick, Maria Isabel Cuevas, Sara Mosquera, Mariana Vásquez, Maria José Correa, Maria Camila Patiño, Pablo Reyes, Lina Maria González Ballesteros

## Abstract

**Background:** Visual and appearance-based social media platforms like TikTok can establish unrealistic beauty standards and self-esteem issues, leading to the development of eating disorders (EDs). This study aims to evaluate the association between TikTok usage and the presence of EDs risk behaviors and body dissatisfaction among Colombian young adults aged between 18 and 25 years.

**Methods:** A cross-sectional descriptive study was conducted via an online survey through snowball sampling with 171 participants. The survey assessed demographic variables, social media use, content consumption, EDs risk behaviors, and body dissatisfaction through validated tools. Non-parametric tests and regression models were used for the data analysis.

**Results:** TikTok users showed significantly higher scores in ED risk behaviors (M = 14.91) and body dissatisfaction (M = 21.9) compared to non-users. Contributory usage, particularly collaborative content creation, was the most associated with increased risk. The multivariate regression model for ED risk explained 3% of the variance, while TikTok use accounted for 30% of the variance in the model for body dissatisfaction.

**Discussion:** Our study found a significant association between TikTok use and the development of ED risk behaviors and body dissatisfaction. These findings align with previous research and highlight the need for interventions that encourage mindful social media consumption.

## Introduction

In recent years, the social network TikTok, created in China in 2016, has become one of the most popular platforms among young people, with 1.8 billion monthly active users (1). The primary content found on this social network consists of short videos covering various topics, including sexualized and appearance-based content, potentially leading users to feel dissatisfied with their bodies and appearance (2). Besides the type of content found on TikTok, other aspects that can influence young people’s behaviors and thoughts include how they use this social network. Consumption patterns range from passive consumption, where the user observes but does not participate, to contributory consumption, where the user creates and contributes content to the social network (3). It has been identified that more active participation within this social network can expose users to issues such as the misuse of personal data, loss of privacy, and problematic use of the platform (4).

Given the influence of social media on new generations, it has been hypothesized that social networks, such as TikTok, could play a role in the onset and maintenance of eating disorders (EDs) (5). Eating disorders, such as anorexia nervosa and bulimia nervosa, are chronic and debilitating conditions characterized by maladaptive eating styles, weight regulation issues, and altered attitudes and perceptions of body image (6). Risk behaviors for EDs are observed in individuals who do not follow culturally normative eating patterns, such as skipping meals, limiting food intake, or following restrictive diets (7). In general, eating behaviors in these cases are associated with inflexible eating rules related to body satisfaction and social security (8).

Considering this, a biopsychosocial model has been proposed to explain the relationship between social media use and the development of self-image problems and maladaptive eating styles (9). This model identifies that the use of social media, which includes predominantly visual and appearance-based content, can set unrealistic beauty standards, leading its users to comparisons and self-esteem issues (10). Additionally, it has been established that the objectification of the female body on social media generates insecurities characterized by constant monitoring of one’s appearance, leading to anxiety and shame. This, in turn, contributes to various mental health issues, such as eating disorders (11).

Therefore, an opportunity arises to investigate how the consumption of popular social media platforms in present times, such as TikTok, can become a risk factor for the development of eating disorders. Consequently, the purpose of this study is to evaluate the association between TikTok usage and the presence of risk behaviors for eating disorders and body dissatisfaction among Colombian young adults aged between 18 and 25 years old. Specifically, the study aims to describe the study population based on TikTok usage, characterize the presence of risk behaviors for eating disorders in relation to TikTok use, assess the prevalence of body dissatisfaction among TikTok users and non-users, compare the frequency and severity of these risk behaviors and body dissatisfaction between TikTok users and non-users, and examine the relationship between the characteristics of TikTok content and the occurrence of risk behaviors for eating disorders and body dissatisfaction in the study population.

## Methods

### Design

This study is a cross-sectional, observational, descriptive study employing non-probabilistic convenience sampling. The sample was collected using a “snowball” sampling approach. The Institutional Ethics and Research Committee of the Pontifical Javeriana University (FM-CIE-0371-24) approved the study.

### Participants

One hundred and seventy-one young adults residing in Colombia were recruited through advertisements posted on social media between May and July of 2024. Participants were included if they were active users of at least one social media platform (defined as using the platform more than three times per week for at least 30 minutes per day), had access to the internet to complete the survey, lived in Colombia at the time of the survey, and were aged 18 to 25 years. Exclusion criteria included having a previous diagnosis of EDs and individuals with neurocognitive disorders and/or cognitive disabilities that would prevent them from completing the survey.

### Procedure

The survey was administered using the REDCap software. After reviewing it, the research team distributed it through email and social media for convenience and requested participants share it further. Eligible participants were also invited to participate in the survey on the campus of Pontificia Universidad Javeriana. The following selection criteria were considered for the participants included in the analysis.

### Measures

#### Demographics

Demographic information was collected from participants, including age, gender, and occupation.

#### Social media consumption patterns

Data on social media usage encompassed the primary social media platform used, the frequency of use over the past seven days, and their engagement with the platform. Engagement was classified into three categories: passive use (viewing or scrolling through content without active interaction), participatory use (interacting with content through likes, comments, or shares), and contributory use (creating and posting original content).

#### Perception of the influence of content on body image in peers

The influence of TikTok content on the perception of body image in peers was assessed using a nominal scale. Participants were asked to evaluate whether they perceived that the content consumed on the platform affected their peers’ body image. Response options included: “Yes, negatively,” “Yes, positively,” “No impact,” and “Not sure.” This measure aimed to capture participants’ views on the broader social impact of TikTok content related to body image within their social circles.

#### Perception of the influence of content on body Image

Participants’ perceptions of how TikTok content influenced their body image were assessed using a nominal scale. They indicated whether they believed the content had a negative impact, a positive impact, no impact, or were unsure. This variable was used to evaluate the subjective impact of social media content on individual body image perceptions.

#### Search for content on diet and weight loss

The frequency with which participants searched for content related to diet and weight loss on TikTok over the past month was measured using an ordinal scale. The scale included the following options: “Never,” “rarely,” “often,” “very often,” and “always.” This variable helped to quantify participants’ engagement with diet and weight loss-related content, providing insights into potential influences on disordered eating behaviors and body image concerns.

#### Body image dissatisfaction

Body dissatisfaction was measured using the Body Dissatisfaction subscale of the Eating Disorder Inventory-2 (EDI-2). This self-report measure consists of nine items evaluated on a six-point Likert scale ranging from 0 (never) to 5 (always). The average score was calculated, with higher scores indicating greater body dissatisfaction. The Colombian version of the EDI-2 has shown high internal consistency, with a Cronbach’s alpha of 0.93 for the overall scale and 0.77 for the Body Dissatisfaction subscale, maintaining reliability despite variations observed in studies from Germany and the Netherlands.

#### Assessment of disordered eating

Disordered eating attitudes were assessed using the Eating Attitudes Test-26 (EAT-26). This self-report questionnaire consists of 26 items scored on a nominal scale with options ranging from “Never” (0) to “Always” (3), except for item 25, which is reverse scored. A higher total score reflects a greater risk of eating disorders. The Colombian validation established a cut-off score of 11 for women and 20 for men, with high sensitivity (100%) and specificity (85.6% for women, 97.8% for men). The internal consistency, measured by Cronbach’s alpha, was 0.92 for women and 0.89 for men, indicating robust reliability in assessing disordered eating symptoms.

### Statistical analysis

Data analysis was performed using Stata software. Initially, a normality test was performed using the Shapiro-Wilk test, revealing that the main variables did not follow a normal distribution (p < 0.05). Hence, non-parametric tests were used to analyze the relationships between variables. Demographic characteristics and social media consumption patterns were described using central tendency and dispersion measures. Differences between TikTok users and non-users were evaluated using Wilcoxon tests for quantitative variables and chi-square tests for categorical variables.

Spearman correlations were developed to explore the association between TikTok use and EDs risk behaviors and body dissatisfaction. Factors that predict EDs risk and body dissatisfaction were evaluated with multivariate linear regression models, controlling for age, gender, occupation, and social media use. The model’s fit was evaluated using the coefficient of determination (R^2^), reported with their respective 95% confidence intervals. Tests were considered significant with a p-value < 0.05.

## Results

A total of 171 participants were included in the analysis. Descriptive statistics for demographic variables can be found in Table 1. Of the total sample, 64 (37.4%) were female and 107 (62.5%) were male. Among male <u>participants</u>, 83.1% reported using the platform, compared to 71.8% of female <u>participants</u>. The age distribution showed asymmetry, with the majority of participants being between 20 and 23 years old, with a mean of 21.48 years (SD = 1.74). When distributed by academic or occupational status, the majority (84.2%) were undergraduate students.

**Table 1:** Sociodemographic characteristics (N = 171)

| Characteristic | Total (%) | TikTok Users (%) | Non-TikTok Users (%) |
| --- | --- | --- | --- |
| Age (Mean ± SD) | 21.48 ± 1.74 | 21.48 ± 1.73 | 21.47 ± 1.80 |
| Gender |  |  |  |
| Male | 107 (62.57%) | 83.18% | 16.82% |
| Female | 64 (37.43%) | 71.88% | 28.13% |
| Occupation |  |  |  |
| Undergraduate Student | 144 (84.21%) | 77.78% | 22.22% |
| Postgraduate Student | 6 (3.51%) | 100.00% | 0.00% |
| Worker (Employed) | 5 (2.92%) | 60.00% | 40.00% |
| Worker and student | 14 (8.19%) | 85.71% | 14.29% |
| No Occupation | 2 (1.17%) | 100.00% | 0.00% |

### Sociodemographic factors and EDs risk

Given that the age variable did not exhibit a normal distribution, a non-parametric Spearman correlation test was conducted to analyze the correlation between ED risk behavior scores and age. The analysis revealed a correlation coefficient of 5.05% between these two variables. Consequently, the null hypothesis of independence between these variables cannot be rejected, indicating insufficient evidence to conclude that age is associated with higher or lower ED risk behavior scores.

Analysis by gender indicated that males exhibited higher ED risk behavior scores, averaging 5 points greater than females. The regression coefficient for females relative to males was statistically significant (CI 95% -10.13 - 1.6). However, the adjusted R^2^ value for the model was only 3.93%, indicating that the model explains a minimal portion of the variance in ED risk behavior scores based on gender alone.

Regarding occupation, postgraduate students had the highest ED risk behavior scores, averaging 35.3, followed by individuals in the “none of the above” category and those studying and working, who averaged 15.75 points. The linear regression model indicated a significant increase in risk behavior scores for postgraduate students (CI 95% 12.6–33.5) and those in the “none of the above” category (CI 95% 1.4–37.1) compared to undergraduate students.

### Social Media Use and EDs risk

The association between ED risk behavior scores and frequency of social media use was assessed using the Spearman test. The results indicated that the null hypothesis of independence between ED risk behavior scores and frequency of use could not be rejected (p = 0.5188). This suggests that there is no statistically significant correlation between frequency of use and ED risk behavior scores within the assessed sample.

The average score of respondents was highest among those who used TikTok, followed by users of other social media platforms like Instagram. However, the relationship between the primary social network used and the score showed a low explanatory power, with an R^2^ value of less than 2%.

Regarding risk behavior related to ED, the reduction in the average risk score was more significant for Instagram users than TikTok users. According to the 95% confidence interval, this reduction was statistically significant.

When comparing the scores of respondents using social media, those who used TikTok had the highest average scores, followed by users of other platforms like Instagram. The relationship between the primary social network used and the scores demonstrated low explanatory power, with an R^2^ value of less than 2%.

Furthermore, the average reduction in the ED risk behavior score was greater for Instagram users than for TikTok users, and this reduction was statistically significant according to the 95% confidence interval.

### TikTok Use and Eating Disorder Risk

TikTok users had an EAT-26 average score of 14.91, while those who did not use the platform scored slightly lower, averaging 9.05. The bivariate analysis showed that the ED risk behavior score was higher on average among participants who used TikTok than those who did not. Variability in scores was more significant among TikTok users. The bivariate model <u>examining</u> the relationship between TikTok usage and risk behavior scores for ED yielded a very low R^2^ value of approximately 3%, suggesting that the model explains only a minimal proportion of the variance in risk behavior scores based on TikTok usage alone.

### TikTok Use and Body Dissatisfaction

Similarly, the average score for body dissatisfaction among TikTok users was 21.9, compared to 15.8 for non-users, as seen on Table 2. The adjusted R^2^ value for the full model assessing body dissatisfaction was 30%, indicating that individuals using TikTok demonstrated higher body dissatisfaction scores than those who did not. Significant predictors of increased scores included age, TikTok usage, commenting on others’ content, and engagement with ED-related content.

**Table 2:** TikTok Use and Risk for Eating Disorders Behaviors.

| Variable | TikTok Users (Mean ± SD) | Non-TikTok Users (Mean ± SD) |
| --- | --- | --- |
| Eating Disorder Risk (EAT-26) | 14.91 ± 14.57 | 9.05 ± 6.60 |
| Body Dissatisfaction (EDI-2) | 21.9 ± 10.33 | 15.8 ± 7.95 |

### Consumption Patterns and EDs risk

The regression model was adjusted with the two categories of passive use of social networks, and it was found that the fit could have been better. However, at a descriptive level, there was an increase of 4 points in ED risk behavior scores for respondents who primarily used TikTok to view or follow others compared to non-users, and viewing friends’ profiles had an average increase of 3.3 points. Regarding participatory use, significant increases in ED risk behavior scores were only noted for individuals sharing content created by others. The analysis of contributive TikTok usage demonstrated a substantial increase in ED risk behavior scores for users participating in duets and collaborative videos, with a rise of 7.82 points. Conversely, individuals producing, editing, and sharing content exhibited a slight, non-statistically significant reduction of -0.15 points. Consumption patterns and EDs risk are shown in Table 3.

**Table 3:**
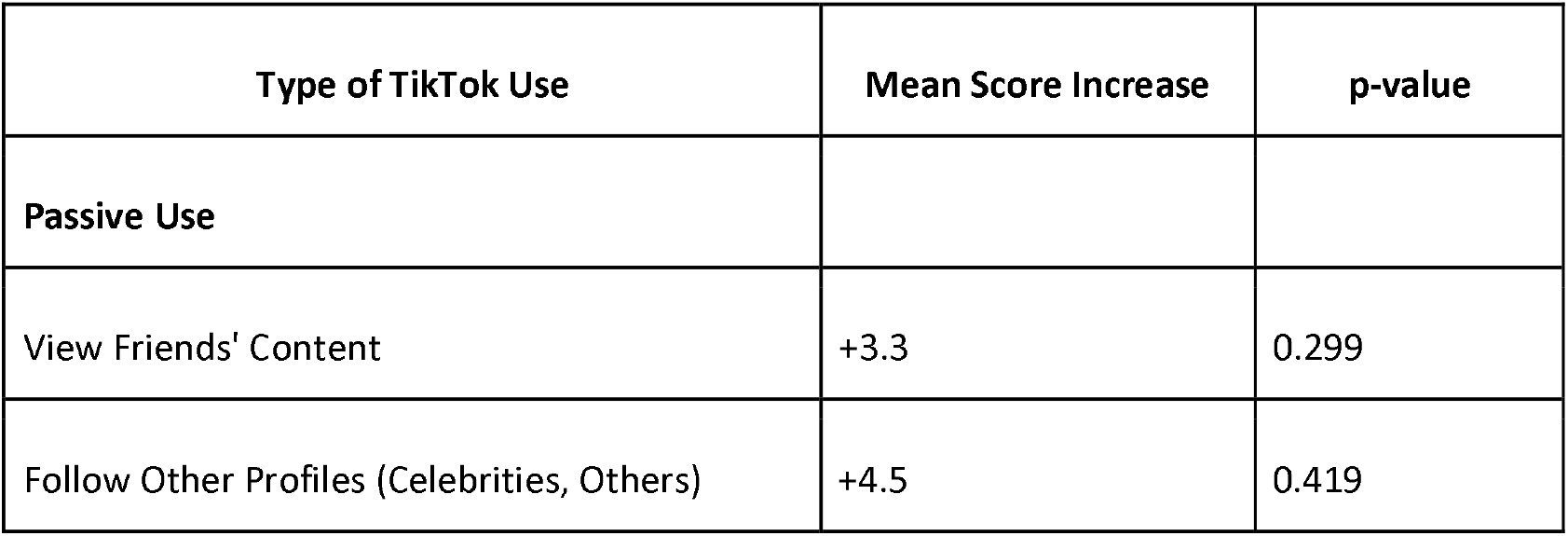
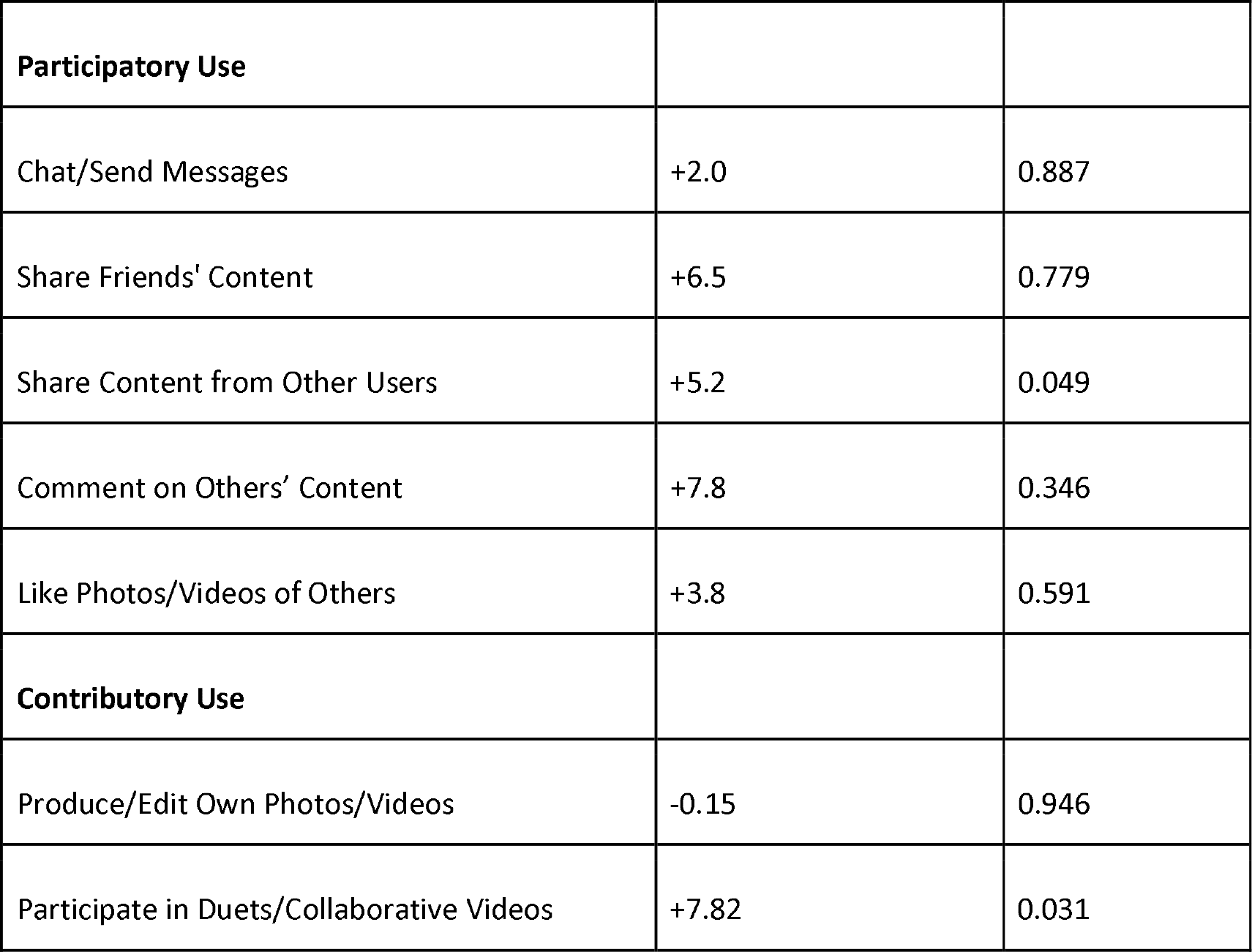
Association Between Types of TikTok Use and Eating Disorder Risk (EAT-26 Scores)

### Other factors associated with EDs risk and body dissatisfaction

The results of the multivariate model for the EAT26 score indicated a good fit, with predictor variables associated with social media use. The model explained 61.83% of the variance. Significant explanatory variables for the increase in the score included using TikTok, frequent and constant engagement with ED-related content, regular content searching, and always imitating behaviors. Other variables showed an increase in the score. Still, they were not statistically significant at the 95% level, including age, being a postgraduate student, using social media to view friends’ photos/videos or follow their profiles, commenting on other users’ photos/videos, liking their content, and frequently imitating behaviors.

Regarding body dissatisfaction, the R^2^ value for the complete model, where the response variable is the body dissatisfaction score, was 30%. Significant variables that increased the score, based on their coefficients and confidence intervals, included age, TikTok usage, commenting on other users’ photos/videos, engaging with ED-related content, and the response of being “not very sure” about the influence of peer content. Other analytical categories showed increases in the score but were not statistically significant at the 95% confidence interval. These included being a postgraduate student, working, using the social media platform categorized as “other,” and using social media to view/follow others’ profiles.

## Discussion

Our study found a significant association between TikTok use and the development of EDs risk behaviors and body dissatisfaction. Our study aligns with previous research on the relationship between TikTok use and body image concerns. It is essential to consider that eating disorders are more prevalent among women despite the majority of participants in this study being men. Despite this, we identified a notable difference in the EAT-26 scores, with TikTok users—regardless of gender—scoring higher on average than non-users. This suggests that TikTok use may exacerbate behaviors associated with eating disorders, although the relationship is complex. While we did find that active TikTok users exhibited a higher risk for eating disorders, the linear regression model revealed a low explanatory value for predicting eating disorder behaviors. This could indicate that TikTok content is not a direct cause or trigger of these disorders. Instead, it may result from an algorithm-driven feedback loop that intensifies exposure to body image issues for those already at risk. Accordingly, previous research has found that social media algorithms can reinforce pre-existing body dissatisfaction by showing appearance-related content to users (12). It has been demonstrated that pro-ana material tends to appear on the user’s bulletin board without actively searching for this content, which has negative self-esteem effects (13).

In contrast, the model for body-image dissatisfaction had a more robust predictive value, explaining 30% of the variability among TikTok users. This supports the notion that while not all TikTok content explicitly promotes disordered eating, the platform’s general focus on idealized body types and physiques can contribute to negative self-perception. The algorithm in general may not exclusively show pro-ana content, but consistent exposure to idealized body standards may still cause dissatisfaction with users’ body image.

From all consumption patterns, contributory TikTok usage was the most associated with increased EDs risk, particularly collaborative content creation. This may be because social media use has been associated with a higher tendency to engage in appearance comparisons, which impacts body dissatisfaction (14). Our findings correlate with previous findings in Colombian females, where Restrepo et al. identified that women who compared their photos on social media with others had a significantly higher risk of developing EDs compared to those who did not (15).

These findings have potential implications for clinical practice in the management of eating disorders among adolescents and young adults. Given TikTok’s pervasive influence on body image, clinicians should consider assessing their patients’ social media habits as part of routine evaluations for eating disorders. Limiting exposure to triggering content could be a valuable tool in therapy, as both the frequency and type of content exposure appear to exacerbate body dissatisfaction (16). Furthermore, healthcare providers should educate patients and their families about the impact of social media and encourage mindful consumption.

To our knowledge, this is one of the first studies to address TikTok use and EDs in Hispanic populations. It addresses a very relevant issue with a potential impact on the mental health of young adults. Another strength is the multifaceted evaluation of social media use. We evaluated the user’s interaction with the content provided, providing insight into the effect of the quality and type of interaction with EDs. We included instruments previously validated on our population, increasing the validity and reliability of our findings. Lastly, multivariate regression models were used to control for several confusion variables, allowing for a more straightforward interpretation of the results.

However, some limitations included our sample size, which may not be large enough to generalize findings and fully represent Colombia’s cultural diversity. The risk of selection bias may also be present since our sample was obtained through online surveys and might have included individuals more likely to use social media. The nature of our study design does not allow for the driving of casualty. While significant associations were found, it is not possible to fully determine if TikTok causes these behaviors or if individuals at a higher risk of developing them are more likely to engage more on social media. Another limitation is self-administered questionnaires, which induce the risk of response bias or social desirability bias. Even though the multivariate analysis controlled for many variables, other unmeasured factors,, such as socioeconomic status or peer influence, might influence the association between social media use and EDs.

Future research should focus on larger, more diverse samples, employing longitudinal studies to understand better the cause-and-effect dynamics between TikTok use and the development of eating disorders. Due to the limitations of our statistical model, which only accounted for a portion of the variances in eating disorders and body image outcomes, it is clear that other factors, such as underlying psychological vulnerabilities or peer influences, play a significant role and deserve further investigation.

## Conclusion

In conclusion, while TikTok content does appear to influence body image perception and may contribute to elevated eating disorder scores, it remains a modifiable factor. This presents an opportunity for interventions to reduce social media use or alter the types of content viewed by young people. By doing so, we can mitigate some of the negative impacts on mental health and promote healthier body image perceptions. However, more research is needed to explore these relationships in greater depth.

## Data Availability

All data produced in the present study are available upon reasonable request to the authors

## References

1. Aslam S. Omnicore. 2023 [cited 2023 May 22]. TikTok by the Numbers (2023): Stats, Demographics & Fun Facts. Available from: https://www.omnicoreagency.com/tiktok-statistics/

2. Bissonette D, Szymanski DM. TikTok use and body dissatisfaction: Examining direct, indirect, and moderated relations. 2022 [cited 2023 May 22]; Available from: 10.1016/j.bodyim.2022.09.006

3. Bucknell Bossen C, Kottasz R. Uses and gratifications sought by pre-adolescent and adolescent TikTok consumers. Young Consumers. 2020 Dec 5;21(4):463–78.

4. Vally Z, D’Souza CG. Abstinence from social media use, subjective well-being, stress, and loneliness. Perspect Psychiatr Care. 2019;55(4):752–9.

5. Holland G, Tiggemann M. A systematic review of the impact of the use of social networking sites on body image and disordered eating outcomes. Vol. 17, Body Image. Elsevier Ltd; 2016. p. 100–10.

6. Kaye W. Neurobiology of Anorexia and Bulimia Nervosa Purdue Ingestive Behavior Research Center Symposium Influences on Eating and Body Weight over the Lifespan: Children and Adolescents. Physiol Behav [Internet]. 2008 Apr 4 [cited 2023 May 22];94(1):121. Available from: /pmc/articles/PMC2601682/

7. Harer KN. Irritable bowel syndrome, disordered eating, and eating disorders. Gastroenterol Hepatol (N Y). 2019;15(5):280–2.

8. Baceviciene M, Jankauskiene R. Associations between Body Appreciation and Disordered Eating in a Large Sample ofll Adolescents. Nutrients. 2020 Mar;12(3).

9. Rodgers RF, Slater A, Gordon CS, McLean SA, Jarman HK, Paxton SJ. A Biopsychosocial Model of Social Media Use and Body Image Concerns, Disorderedll Eating, and Muscle-Building Behaviors among Adolescent Girls and Boys. J Youth Adolesc. 2020 Feb;49(2):399–409.

10. O’Brien KS, Caputi P, Minto R, Peoples G, Hooper C, Kell S, et al. Upward and downward physical appearance comparisons: development of scales andll examination of predictive qualities. Body Image. 2009 Jun;6(3):201–6.

11. Harper BJ, Tiggemann M. The Effect of Thin Ideal Media Images on Women’s Self-Objectification, Mood, and Body Image. Sex Roles. 2008;58:649–57.

12. Harriger JA, Evans JA, Thompson JK, Tylka TL. The dangers of the rabbit hole: Reflections on social media as a portal into a distorted world of edited bodies and eating disorder risk and the role of algorithms. Body Image [Internet]. 2022;41:292–7. Available from: https://www.sciencedirect.com/science/article/pii/S1740144522000638

13. Pruccoli J, De Rosa M, Chiasso L, Perrone A, Parmeggiani A. The use of TikTok among children and adolescents with Eating Disorders: experience in a third-level public Italian center during the SARS-CoV-2 pandemic. Ital J Pediatr. 2022 Jul;48(1):138.

14. Rodgers RF, Slater A, Gordon CS, McLean SA, Jarman HK, Paxton SJ. A Biopsychosocial Model of Social Media Use and Body Image Concerns, Disordered Eating, and Muscle-Building Behaviors among Adolescent Girls and Boys. J Youth Adolesc. 2020 Feb;49(2):399–409.

15. Restrepo JE, Castañeda T. Riesgo de trastorno de la conducta alimentaria y uso de redes sociales en usuarias de gimnasios de la ciudad de Medellín, Colombia. Rev Colomb Psiquiatr [Internet]. 2020; 49(3):162–9. Available from: http://www.scielo.org.co/scielo.php?script=sci_arttext&pid=S0034-74502020000300162&lng=en.

16. Sanzari CM, Gorrell S, Anderson LM, Reilly EE, Niemiec MA, Orloff NC, et al. The impact of social media use on body image and disordered eating behaviors: Content matters more than duration of exposure. Eat Behav. 2023 Apr;49:101722.

